# Enhancing system empathy within a UK Emergency Department: A feasibility interprofessional priority setting exercise

**DOI:** 10.1101/2024.04.15.24305826

**Authors:** Jeremy Howick, Andy Ward, Charlotte Grantham, Amber Bennett-Weston

## Abstract

**Background:** System-level barriers inhibit empathy in healthcare, and this can harm patients and practitioners. The barriers include burnout-inducing administrative workloads, burdensome protocols, lack of wellbeing spaces, un-empathic leadership, and not emphasising empathy as an institutional value. A workshop aimed at enhancing empathic systems was successfully delivered in Canada but has not been tested in the UK National Health Service (NHS) setting.

**Aim:** To test the feasibility of an empathic systems workshop within the UK NHS setting.

**Methods:** We conducted an interprofessional group of an emergency department (ED). We used a modified nominal group technique to prioritise actions to enhance empathy in the ED system. Satisfaction with the workshop and confidence that the workshop would lead to change were measured on a 10-point Likert scale.

**Results:** Twenty-eight participants representing the following stakeholder groups attended the workshop: leaders, consultants, nurses, security, and porters. The group agreed to generating a better wellbeing action plan and implementing an effective secondary triage system. Seventy-three percent (73%) rated their satisfaction with the workshop as 8 or higher out of ten, and 63% reported being confident that the workshop would lead to positive change.

**Limitations:** A doctors strike limited the range of stakeholders who were able to attend, and long-term follow up was not conducted.

**Conclusions:** Participants in a UK setting were satisfied with a previously developed system empathy workshop and reported being confident that it would lead to positive change. Participants were able to prioritise changes that would improve system empathy and were confident that the changes would be effective.

## 1 INTRODUCTION

The Ockenden, Francis, and Kirkup reports into the fatal tragedies within NHS hospitals concluded that lack of empathy (often defined as caring action with interpersonal understanding^1^) contributed to hundreds of avoidable deaths.^2–4^ Similar findings have been reported in the US.^5^ Lack of empathy has been shown to increase the risk of medical errors,^6^ patient complaints,^7^ and medico-legal risks.^8^ Meanwhile, patients who are treated by empathic practitioners have better outcomes, ranging from satisfaction with care to pain and quality of life.^8^ Contrary to what is sometimes assumed, empathy can be taught.^9^

Empathy in healthcare has often been discussed in the context of “relational empathy,” between patients and practitioners.^10^ However, busy and overworked practitioners may not have time for empathy training (or, if they do take time, the increased workload could add stress to an already stressful job).^11^ Stress, in turn, stifles the ability to relate, understand, and empathise.^12^ Practitioners also work with often inefficient and confusing electronic health records systems^13–15^ that take up half of their time,^16^ managerialism,^17^ and a culture where bullying is common^18^ and empathy is lacking.^19^ These problems are exacerbated by increasingly dissatisfied patients^20^ who may have been greeted by an unwelcoming receptionist^21^ and made to wait for their appointment for an unacceptably long time.

In short, healthcare practitioners operate within a system which can either foster or dampen empathy between themselves and their patients.^22^ For empathy to thrive, the system must be conducive to it.^22^

To promote empathic systems, an interprofessional workshop was conducted at a hospital in Canada.^23^ That workshop led to changes including more welcoming spaces and linked patient appointments.^24^ However, that workshop has not been adopted for a UK setting.

### 1.1 Aims and objectives

The overall aim of this workshop was to test whether a system empathy workshop conducted in Canada was feasible in an UK National Health Service (NHS) setting. The aims of the original workshop^23^ were (1) to identify barriers and facilitators to empathic care within a specific system; (2) to identify and prioritise changes that would improve the degree of empathy within the system; and (3) to develop and commit to a strategy for making the prioritised changes. Our workshop had the same aims.

To test whether the workshop was feasible, we collected feedback regarding the satisfaction of the workshop, confidence regarding whether the workshop would lead to positive change, and what participants would improve about the workshop.

## 2 METHODS

Institutional review board approval was not obtained as the study was a report of an educational leadership workshop. We used relevant items from the Reporting guideline for priority setting of health research with stakeholders (REPRISE) to report this study.^25^ See supplementary Table 1 for a completed REPRISE checklist.

### 2.1 Underpinning theories of change

The workshop was underpinned by the nominal group technique, deep interdisciplinarity and stakeholder engagement, blue sky thinking, prioritisation, and urgency. While none of these are unique individually, we are not aware of them being combined.

#### 2.1.1 Nominal group technique

The nominal group technique is recommended by REPRISE as an efficient way to identify and generate consensus on priorities in healthcare.^26^ ^27^ It involves structured group brainstorming that encourages participation from all participants.^28^ Quantitative and qualitative data are combined by having participants write down their ideas, voting on the ideas they believe are best.

#### 2.1.2 Deep interdisciplinarity and stakeholder engagement

Organisational change appears to be more effective when stakeholders (including all employees) are the agents of change.^29^ This is contrasted with the common top-down initiatives from central bodies such as the NHS or government. Top-down initiatives reflect a managerial leadership model which can exacerbate disunity,^30^ increase stress,^31^ and reduce empathy.^2^ ^3^ Despite interprofessionalism being promoted,^32^ the involvement of a wide range of stakeholders has rarely been adopted in priority setting exercises.^33^ Additionally, insofar as interprofessionalism is adopted, it often excludes ancillary staff and managers.^34^ This is a mistake because receptionists,^35^ ^36^ cleaners, ^37^ porters, security guards,^38–40^ and others can influence patient experience and empathy,^41^ ^42^ as well as other system-level changes.^43^ Leading managers are important stakeholders for ensuring credibility and acceptability of uptake.^44^ ^45^ These leaders have more power to implement changes,^46^ and involving them in a diverse group of stakeholders helps to break down the “us versus them” barrier to effective change.^47^ Finally, especially in the healthcare setting, including patient perspectives and voices is considered a duty as well as a useful way to generate suggestions that will improve patient care.^48–50^ While we hoped to have patient representatives in our workshop, the context prevented this (see limitations).

#### 2.1.3 Blue sky thinking

Leaders and researchers sometimes report that they lack the time, space, or inclination to generate creative solutions to problems.^51^ Blue sky thinking frees people from normal constraints to encourage creativity, and helps shift the focus from problems towards solutions.^52^

#### 2.1.4 Prioritisation and commitment

The adage that if everything is important, nothing is important holds true for effective organisational change.^53^ Yet, calls for organisational change often include dozens (or more) quality improvement recommendations. To wit, the 2013 Francis Report contained 290 recommendations,^3^ and nearly a decade later, the Ockenden^2^ report contained 15 “immediate and essential” recommendations. Unsurprisingly, there is a lack of evidence that any of the recommendations have been successfully implemented.^54^ While providing a long list of recommendations may appear comprehensive, without clear priorities it is hard to know where to start and efforts become diluted to the point of ineffectiveness. This, in turn, can create frustration and a sense of helplessness.

Failure to prioritise also suggests lack of rigour. From a long list of recommendations, some will have a bigger effect than others. Indeed, some may leverage large and far-reaching effects (“power law” initiatives).^55^ By contrast, choosing just one, two, or three priorities that stakeholders believe will have the biggest effect focuses hearts and minds on making progress.^56^ In addition to prioritising, it is important to get stakeholders to commit. Research suggests that people are more likely to remain committed to something if they explicitly commit to it, for example by writing it down on paper.^57^

#### 2.1.5 Creating a sense of urgency

Having long-term visions for major initiatives is good. However, once the priorities have been set and there is a plan in place, it is important to set an ambitious timeline that prevents people from being able to procrastinate. This timeframe is often between 6 and 12 months.^29^ ^53^ Another reason for using short timelines is that it reduces the risk of unanticipated events that can derail the project.^55^ These events can include anything from key leaders changing jobs, to pandemics. The shorter the timeframe, the less risk there is of these events derailing the project.

### 2.2 Details of workshop

#### 2.2.1 Participants

The participants worked in the Emergency Department (ED) within the University Hospitals of Leicester (UHL). The context of this workshop involved junior doctor strikes and ensuing pressure on all staff, which limited the ability to choose a completely representative sample of stakeholders. Participants were therefore a convenience sample chosen by the lead of the ED.

#### 2.2.2 Healthcare setting

We chose the ED because a patient’s journey through the hospital often begins with the emergency department. At a time of high stress and emotion for the patient and their loved ones, this chaotic and overwhelming environment can set a negative first impression of their care to come. Several factors that impact the patient’s experience of care in ED include overcrowding, lack of privacy, busy and overworked practitioners, all of which can result in dissatisfied patients.^58^ This makes EDs places where an impact can be made.^59^

#### 2.2.3 Workshop setting

The workshop was held at a central location with convenient access for attendees. It was conducted during working hours as part of attendees’ allocated training time, and refreshments and lunch were provided.

#### 2.2.4 Workshop conduct

Two experienced facilitators (AW, JH) conducted the session, and participants were seated at tables in groups of 5-6 people. Participants were made aware that the scope of the workshop was “system empathy,” for which a definition was provided.^24^ Systemic factors were explained to include external environmental factors, organisational factors, physical environment, job factors, and individual characteristics.^24^ The small groups then participated in five exercises (see Table 1).

1. **System level barriers**. In the first exercise participants were asked to consider the system-level barriers to empathic healthcare that they faced.
2. **Empathic healthcare in an ideal world**. The group then considered the system-level facilitators of empathic healthcare. To encourage “blue sky thinking,” the groups were asked to imagine the ideal situation in their department – a situation where they were given access to unlimited resources and power to make their department more empathic. Each table was given 15 minutes and tasked to produce a poster to summarise their ideas with a prize for the best.
3. **Empathic healthcare in your setting**. Next, participants considered how to overcome barriers and facilitate empathic healthcare in their team and setting. The first exercise required them to think practically about actions they could take, being ambitious but pragmatic. They were asked to write their individual ideas onto sticky notes and add them to a flipchart in the centre of their table. Participants were then encouraged to read the other groups’ ideas and copy any that they thought should be added to their own flipchart.
4. **Prioritisation**. A modified nominal group technique was used to prioritise the ideas on each table with the aim of narrowing down to a small number (up to three) priorities that could be achieved within the next six to 12 months. Each participant was given three yellow stickers to add to the sticky notes with their top-rated ideas. In the next round of voting, everyone was given a single green sticker to apply to their top priority. Interventions on each table with the greatest number of green stickers were chosen for implementation.
5. **Implementation plans**. Finally, each small group developed plans for how to implement, encourage, and measure progress for the chosen interventions. These were recorded on SMART Action Plan templates (provided) with a named person made responsible for implementing each. The action plans were then shared and discussed with the larger group.

**Table 1.** Structure and timings of the workshop.

| **Activity** | **Time allotted** |
| --- | --- |
| Introductions, aims and objectives | 15 minutes |
| Icebreaker Exercise | 20 minutes |
| Presentation – The evidence for empathic healthcare and empathic systems | 30 minutes |
| Group exercise – System Barriers to empathic healthcare<br>*Work in groups of 2-3 for 10 minutes, share with rest of table, share with larger group* | 30 minutes |
| **Active Break** | **30 minutes** |
| Group Exercise – Empathic healthcare in an ideal world<br>*Work in groups of 2-3 for 10 minutes, share with rest of table, share with larger group* | 60 minutes |
| Self-empathy<br>*Guided meditation (whole group – optional)* | 20 minutes |
| **Active Lunchbreak** | **60 minutes** |
| Group Exercise – How to overcome barriers and facilitate empathic healthcare in your setting<br>*15 minutes generating ideas, 15 minutes circulating, reading, stealing ideas* | 30 minutes |
| Group Exercise – Prioritisation<br>*Nominal voting* | 30 minutes |
| **Short break** | **15 minutes** |
| Action planning<br>*Creating, sharing and discussing SMART plans* | 50 minutes |
| Postcard exercise | 10 minutes |
| Close and summary | 5 minutes |

At the end, participants were also provided with postcards where they could write what they were willing to commit to do individually to make the department more empathic. The postcards were collected and then mailed to the participants a week later as a reminder of their commitment. Throughout the process, participant engagement was cultivated with an icebreaking exercise, a caption competition (with a prize), short humorous video clips, active breaks and an optional meditation session.

### 2.3 Data Collection

Data collection was done by a note taker with a medical background. This data was supported by transcribed pictures of the flipcharts used by the participants to write their ideas.

### 2.4 Analysis

A recent study used the human factors approach to specify the features of an empathic healthcare system.^60^ They divided the factors into external environmental factors (such as the way in which healthcare practitioners are reimbursed), organisational factors (such as empathic leadership), physical and technical environment (whether there are healing spaces), job factors (such as sensible workload), and individual characteristics (such as wellbeing). We used this framework to categorise the facilitators and barriers (see Table 2).

**Table 2.**
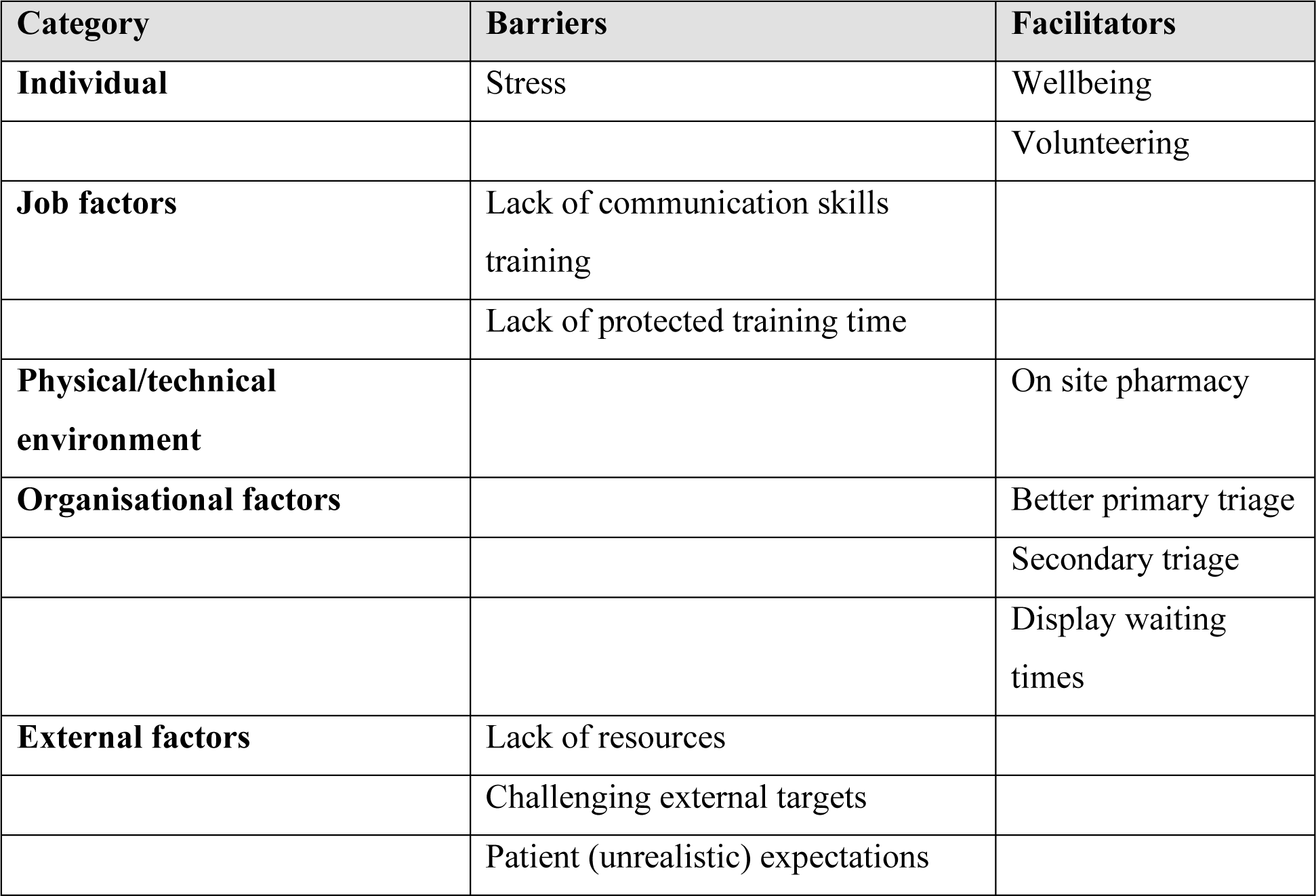
Summary of barriers and facilitators.

### 2.5 Evaluation

Participants were asked to complete a short online survey at the end of the workshop to evaluate their satisfaction with the session and their confidence that it would improve teamworking in the ED. The survey included an adapted ‘Friends and Family Test’ question, which asked participants whether they would recommend the ED as a place to work *if* they were to implement their learning from the workshop.

## 3 RESULTS

### 3.1 Participants Characteristics

Twenty-eight participants from the following groups took part: consultants (n=1), nurses (n=20), administrators (n=5), and porters (n=2).

### 3.2 Barriers to Empathic Care

Participants listed six main barriers to empathy in the ED setting: stress, communication, patient expectations, lack of resources, lack of empathy training, and challenging external targets (see Table 2).

#### Stress

All participants agreed that the ED was an intense environment to work in. They also noted that anxiety and stress left little emotional “bandwidth” for clinical and non-clinical staff to communicate empathically with patients.

#### Communication skills training

Whilst some professional groups had received communication skills training, this was not universal, and it was felt that all staff at all levels could benefit. Communication with patients, colleagues and professionals outside the department were identified as areas for development.

#### Patients’ (unrealistic) expectations

Many participants in the workshop noted that high public expectations put added pressure and emotional strain on a system already working beyond capacity. The increase in media coverage of NHS services and social media attention over the years could be a contributing factor.

#### Lack of resources

The participants noted a lack of human resources as well as infrastructure resources (including space and equipment). This led to overcrowding and a comparatively tumultuous environment that made empathy between patients and both clinical and non-clinical staff difficult.

#### Lack of protected training time

Whereas the benefits of training (including empathy training) were acknowledged, it was noted that staff did not have time to attend teaching or training opportunities. This led to a feeling of being undervalued and underinvested. This was especially acute for trainees. The feeling of being undervalued led to lower staff satisfaction, which participants found stood in the way of patient-practitioner empathy.

#### Challenging external targets

Managerial pressures to reach targets such as wait time targets. These targets contributed to all the barriers listed above, including stress, poor communication, the feeling that there is not enough resource, and insufficient time for training.

### 3.3 Empathic care in an ideal world

For factors that could improve system empathy, unrestricted by resources, workshop participants listed the following six: on-site pharmacy, displaying waiting times, triage of frequent attenders, secondary triage, a wellbeing group, and volunteering.

#### On-site pharmacy

It was believed that having an on-site pharmacy would lead to a smoother discharge and improved adherence, which might reduce the number of avoidable repeat visits due to failure to take medication.

#### Display waiting times

It was claimed that patients often complain about not knowing how long they might have to wait. Dissatisfied patients, in turn, could become “difficult” and less easy to communicate with (empathically).

#### Appropriate triage of people who attend the department frequently

Participants specified a disproportionate number of ED visits were from frequent attenders. This patient cohort often experiences multiple disadvantages and have complex social needs that are challenging or sometimes inappropriate to address in an acute care setting. It was believed that if this cohort could be triaged more appropriately and signposted to appropriate community services, they would receive care that would help to avoid acute admission. For example, people experiencing homelessness with complex social needs could be directed to local charities for support with housing, food, clothing and addiction services if this was more appropriate for their presenting issues.

#### Secondary triage

Some participants suggested that secondary triage would be appropriate for reducing waiting times.

#### Improved wellbeing

It was believed that wellbeing could be improved with a better wellbeing programme for both clinical and non-clinical staff. It was agreed that there were various wellbeing initiatives in the hospital, but staff were not always aware of them and, insofar as they were aware, found them to be lacking.

#### Volunteering

Several participants made the others aware of the health benefits of volunteering and proposed that more opportunities to volunteer would be beneficial.

### 3.4 Priorities

Following the prioritisation exercise, two initiatives were prioritised, and participants agreed to take them forward: a wellbeing subgroup, and secondary triage. A subgroup of participants agreed to meet to consolidate what was offered by UHL and how it could be better communicated. A senior consultant who attended the workshop agreed to research the most effective and evidence-based ways to introduce secondary triage.

### 3.5 Analysis

The barriers and facilitators fell into a range of categories of system level factors that affect empathy. Challenging external targets, lack of resources, and patients’ (unrealistic) expectations are *external environmental factors*, failure to provide empathic communication skills training and lack of protected training time are *job factors* and stress an *individual factor*. Of the facilitators, three (displaying waiting times, triage of frequent attenders, and secondary triage) could be classified as *organisational factors*, one as part of the *physical and technical* environment (on-site pharmacy), and two (wellbeing, volunteering) as *individual characteristics*. Despite noting that several external environmental factors and job factors were barriers, none of the proposed facilitators mentioned these. This could be because many of the external environmental factors are out of the participants’ control. Also, whereas none of the barriers listed were related to the physical and technical environment, many of the proposals were. The mismatch could have been due to the limitations, such as potential over-representation from senior consultants.

### 3.6 Survey Results

Nineteen out of the 28 participants (68%) filled in the feedback survey and consented to have their responses published (Table 3). Most (14/19, 73%) of the participants rated their satisfaction with the workshop at 8 or higher out of 10. Twelve (63%) reported being confident that the system and teamwork would improve because of the workshop. All (100%) of participants answered ‘yes’ when asked if they would recommend the ED to their friends and family as a place to work, if they were to successfully implement their learning from the workshop.

**Table 3.**
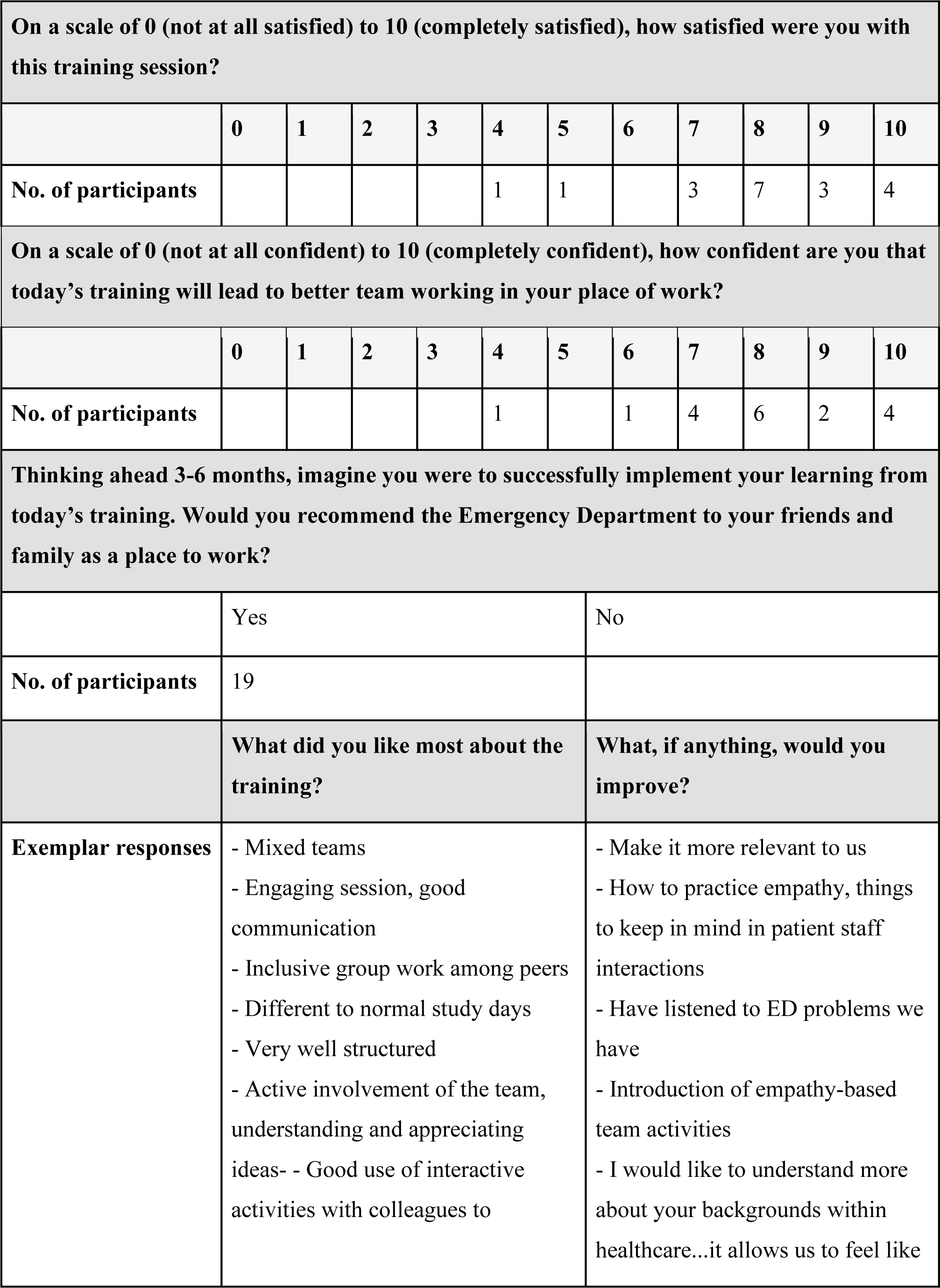
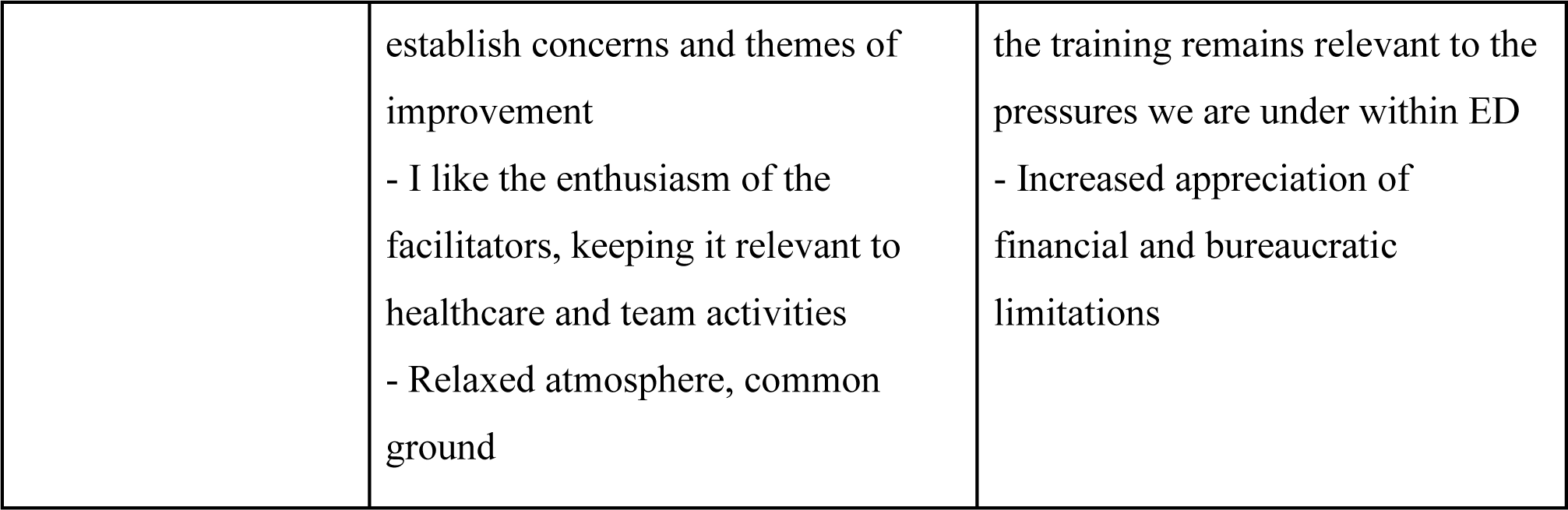
Survey results.

| On a scale of 0 (not at all satisfied) to 10 (completely satisfied), how satisfied were you with this training session? |  |  |  |  |  |  |  |  |  |  |  |
| --- | --- | --- | --- | --- | --- | --- | --- | --- | --- | --- | --- |
|  | 0 | 1 | 2 | 3 | 4 | 5 | 6 | 7 | 8 | 9 | 10 |
| No. of participants |  |  |  |  | 1 | 1 |  | 3 | 7 | 3 | 4 |
| On a scale of 0 (not at all confident) to 10 (completely confident), how confident are you that today's training will lead to better team working in your place of work? |  |  |  |  |  |  |  |  |  |  |  |
|  | 0 | 1 | 2 | 3 | 4 | 5 | 6 | 7 | 8 | 9 | 10 |
| No. of participants |  |  |  |  | 1 |  | 1 | 4 | 6 | 2 | 4 |
| Thinking ahead 3-6 months, imagine you were to successfully implement your learning from today's training. Would you recommend the Emergency Department to your friends and family as a place to work? |  |  |  |  |  |  |  |  |  |  |  |
|  | Yes |  |  |  |  |  | No |  |  |  |  |
| No. of participants | 19 |  |  |  |  |  |  |  |  |  |  |
|  | What did you like most about the training? |  |  |  |  |  | What, if anything, would you improve? |  |  |  |  |
| Exemplar responses | - Mixed teams<br>- Engaging session, good communication<br>- Inclusive group work among peers<br>- Different to normal study days<br>- Very well structured<br>- Active involvement of the team, understanding and appreciating ideas-<br>- Good use of interactive activities with colleagues to |  |  |  |  |  | - Make it more relevant to us<br>- How to practice empathy, things to keep in mind in patient staff interactions<br>- Have listened to ED problems we have<br>- Introduction of empathy-based team activities<br>- I would like to understand more about your backgrounds within healthcare...it allows us to feel like |  |  |  |  |

|  |  |  |
| --- | --- | --- |
|  | establish concerns and themes of improvement<br>- I like the enthusiasm of the facilitators, keeping it relevant to healthcare and team activities<br>- Relaxed atmosphere, common ground | the training remains relevant to the pressures we are under within ED<br>- Increased appreciation of financial and bureaucratic limitations |

In the written feedback, there was an overall theme of appreciation for the opportunity to interact with colleagues in different roles from across the ED. Participants explained that this helped to establish “…concerns and themes of improvement” required to enhance team working and create a more empathic system. Participants also valued the “enthusiasm” of the facilitators, by whom they felt empowered to make changes in their workplace. For future sessions, it was suggested that greater consideration could be given to the specific challenges faced by staff in the ED. Relatedly, one participant expressed a need for “increased appreciation of financial and bureaucratic limitations.” Anecdotally, from participants’ verbal feedback during the course, the humorous videos and energetic demeanour of facilitators helped with engagement.

## 4 DISCUSSION

### 4.1 Summary of findings

A successful and well-received workshop was delivered to members of a large hospital department in the UK that has the potential to enhance the empathy of the system in which they work.

### 4.2 Comparison with other evidence

A similar workshop was conducted at a children’s hospital in Montreal, Canada.^23^ Both workshops were similar in that they were be feasible and enjoyed by participants. Also, some of the barriers to empathy were shared (stress, lack of communication skills training). However, whereas participants in this workshop listed (unrealistic) patients’ expectations, lack of protected training time, and challenging external targets, the participants in the Canadian workshop listed other barriers (inefficient technology, paperwork, complexity of patients, need to train new doctors, cold and unwelcoming infrastructure, and failure to acknowledge the importance of empathy). Relatedly, while several facilitators were shared (the need for staff wellbeing support), the participants in the Canadian workshop mentioned other facilitators that were not mentioned in the workshop described above (embedding empathy as an institutional goal, offering incentives for empathic care, brighter and more open spaces, additional time for trainees, more interdisciplinarity, more admin staff, and patient-centred technology). These differences are to be expected given the different contexts. In fact, a strength of the workshop is that stakeholders from organisations develop priorities that are specific to their setting. In this way, variation is expected, and most likely to be an advantage as it evidences that the participants have generated a bespoke, context-specific strategy.

All the barriers identified by our participants have been noted in the literature. Specifically, stress and burnout are widespread and increasing,^61^ and a likely barrier to empathy.^62^ Patients’ expectations have changed, and some have noted that patients’ rights may currently be over-emphasised in comparison with patient responsibilities.^24^ Unsurprisingly, a study showed that lack of resources diminished the quality of care,^63^ although adding resources to a system that does not manage resources well has will not solve the problem.^64^ and could even make it worse.^65^ Finally, external targets such as the Quality Outcomes Framework (QOF) is often listed as something that gets in the way of practitioners focusing on patients,^66^ ^67^ although it has been shown to improve many patient outcomes.^68^

Relatedly, the workshop participants’ suggestion to include pharmacies within the department is supported by evidence. For example, while the American College of Emergency Physicians recommends that clinical pharmacists be located within emergency departments,^69^ the recommendation is often not followed, and the evidence that in house clinical pharmacists improve wait times are constantly reported amongst literature as one of the greatest factors impacting their ED experience.^58^ One study found that time spent waiting before a consultation negatively impacts patient-reported empathy scores.^70^ Relatedly, patients perceive displaying wait times as positive and needed.^71^ Triage of frequent attenders has also been noted as important. Roughly one in five of all ED visits are from frequent attenders,^72^ and this patient group often feels that healthcare professionals lack empathy and downplay their needs. A lack of empathy increased frequent attenders’ stress levels and reduced patient satisfaction.^73^

The patient benefits of improving triage have been noted in a systematic review of 50 primary studies,^74^ and triage is often susceptible to improvement.^75^ Hence, the prioritisation of secondary triage seems likely to bear fruit.

Volunteering within a healthcare setting has several benefits for the mental and physical health of the volunteer. A volunteering scheme for medical and nursing students in US has shown an increase in empathy levels, the development of new perspectives on the patient experience, improved confidence in patient communication, a stronger professional identity and a rewarding experience.^76^ The value of volunteering is reflected in healthcare policies such as the NHS Long-term plan and NHS workforce plan. Relatedly, the participants’ suggestion to focus on improved wellbeing is reflected in key guidance documents. For example, the Royal College of Emergency Medicine recently released their 2024 key recommendations for Retention and Workforce Wellbeing in Emergency Care.^77^ This includes recommendations on physical environment, protected study time, culture change to more compassionate leadership, inter-professional valuing and respect.

### 4.3 Strengths and limitations

Strengths of our research included collaboration between academia and healthcare practitioners, and “deep interdisciplinarity.” This was achieved by having a multi-disciplinary team. Our workshop and study also had some limitations. While the individuals within the workshop were representatives from a variety of stakeholders, successful implementation of the recommended changes would be further promoted by an even wider participation and targeted groups. In our workshop, there was only one senior consultant and no patient representatives. Future iterations of this workshop should involve more targeted selection of participants from a wider range of stakeholders. Another weakness is that long-term outcomes were not measured.

### 4.4 Recommendations for future research and practice

The workshop must be modified to account for the feedback received. Specifically, empathy-focused activities could be introduced, and facilitators with more experience in the particular setting would improve the credibility and buy-in from participants.

Further research is required in several areas. First, long-term outcomes of the system empathy workshop must be measured. Second, the theory underpinning system empathy should be further explored, perhaps informed by reverse engineering this training. Relatedly, a framework for system empathy should be developed to facilitate measuring and implementing system empathy at other institutions.

### 4.5 Conclusion

Empathic encounters between patients and practitioners in the hospital setting should not be experienced as a luck of the draw, but something that patients can count on to have throughout their journey. The most needed initiatives to provide systemic empathy at our tertiary emergency department were increasing the number of welcome volunteers, introducing an on-site pharmacy and implementing a secondary triage service.

#### Grant information

JH, AW, and ABW are supported by the Stoneygate Trust (grant number not applicable). The funder had no role in the conceptualisation, design, data collection, data analysis, decision to publish, or preparation of the manuscript.

## Data availability statement

Data related to this research is included in this publication. Please contact the corresponding author if additional information is required.

## Acknowledgements

Catherine Eyres (CE) and Catherine Nunn (CN) organised the workshop. CE collected some data and proofread the article.

## Ethics

Institutional review board approval was not obtained as the study was a report of an educational leadership workshop.

