## Supplementary Table 1 for "Enhancing system empathy within a UK Emergency Department: A feasibility interprofessional priority setting exercise"

**Supplementary Table 1. REPRISE Checklist**

| **No.** | **Item** | **Descriptor and/or examples** | **Page No.** |
| --- | --- | --- | --- |
| A | Context and scope |  |  |
| 1 | Define geographical scope | Global, regional, national, city, local area, institutional/organizational level, health service | 6 |
| 2 | Define health area, field, focus | Disease or condition specific, interventions, healthcare delivery, health system | 6 |
| 3 | Define the intended beneficiaries | This may include the general population or a specific population based on demographic (age, gender), clinical (disease, condition), or other characteristics who may benefit from the research | 4 |
| 4 | Define the target audience of the priorities | Policy makers, funders, researchers, industry or others who have the potential to implement the priorities identified | 6 |
| 5 | Identify the research area | Public health, health services research, clinical research, basic science | 3-4 |
| 6 | Identify the type of research questions | Etiology, diagnosis, prevention, treatment (interventions), prognosis, health services, psychosocial, behavioral and social science, economic evaluation, implementation; this may not be pre-defined | 3-4 |
| 7 | Define the time frame | Interim, short-term, long-term priorities, plans to revise and update | 7 |
| B | Governance and team |  |  |
| 8 | Describe the selection and structure of the leadership and management team | Those responsible for initiating, developing, and guiding the process for priority setting, and examples of structures include; Steering Committee, Advisory Group, Technical Experts | 6-7 |
| 9 | Describe the characteristics of the team | Stakeholder group or role, institutional affiliations, country or region, demographics (e.g. age sex), discipline, experience, expertise | 6-7 |
| 10 | Describe any training or experience relevant to conducting priority setting | Consultants or advisors, members with experience or skills relevant to the conducting priority-setting e.g. qualitative methods, surveys, facilitation | 7 |
| C | Framework for priority setting |  |  |
| 11 | State the framework used (if any) | James Lind Alliance, COHRED, CHNRI, Dialogue Model, no framework (general research priority setting) | N/A |
| D | Stakeholders or participants |  |  |
| 12 | Define the inclusion criteria for stakeholders involved in priority-setting | Patients, caregivers, general community, health professionals, researchers, policy makers, non-governmental organizations, government, industry; specific groups including vulnerable and marginalized populations | 6 |
| 13 | State the strategy or method for identifying and engaging stakeholders | Partnership with organizations, social media, recruitment through hospitals | 6 |
| 14 | Indicate the number of participants and/or organizations involved | Number of individuals and organizations, include number by stakeholder group | 9 |
| 15 | Describe the characteristics of stakeholders | Stakeholder group, demographic characteristics, areas of interest and expertise, discipline, affiliations | 9 |
| 16 | State if reimbursement for participation was provided | Cash, vouchers, certificates, acknowledgement; what purpose e.g. travel, accommodation, honorarium | 7 |
| E | Identification and collection of research priorities |  |  |
| 17 | Describe methods for collecting initial priorities | Methods e.g. Delphi survey, surveys, nominal group technique, interviews, focus groups, meetings, workshops; prioritization e.g. voting, ranking; mode e.g. face-to-face, online; may be informed by evidence e.g. systematic reviews, reviews of guidelines/other documents, health technology assessment | 7 |
| 18 | Describe methods for collating and categorizing priorities | Taxonomy or other framework used to organize, summarise, and aggregate topics or questions | 7-8 |
| 19 | Describe methods and reasons for modifying (removing, adding, reframing) priorities | Based on scope, clarity, definition, duplication, other criteria | 7-8 |
| 20 | Describe methods for refining or translating priorities into research topics or questions | Reviewed by Steering Committee or project team | N/A |
| 21 | Describe methods for checking whether research questions or topics have been answered | Systematic reviews, evidence mapping, consultation with experts | N/A |
| 22 | Describe number of research questions or topics | Number of priorities at each stage of the process | 7-8, 11 |
| F | Prioritization of research topics/questions |  |  |
| 23 | Describe methods and criteria for prioritizing research topics or questions | Methods e.g. Delphi survey, surveys, nominal group technique, interviews, focus groups, meetings, workshops;  Prioritization e.g. voting, ranking;  Mode e.g. face-to-face, online;  Criteria e.g. need, feasibility, novelty, equity | 7-8 |
| 24 | State the method or threshold for excluding research topics/questions | Thresholds for ranking scores, proportions, votes; other criteria | 7-8 |
| G | Output |  |  |
| 25 | State the approach to formulating the research priorities | Area, topic, questions, PICO (population, intervention, comparator, outcome) | 7-8 |
| H | Evaluation and feedback |  |  |
| 26 | Describe how the process of prioritization was evaluated | Survey, workshop | 8 |
| 27 | Describe how priorities were fed back to stakeholders and/or to the public; and how feedback (if received) was addressed and integrated | Public meetings or workshop, newsletters, website, email, online presentations | 8 |
| I | Implementation |  |  |
| 28 | Outline the strategy or action plans for implementing priorities | Communication with target audience, via policies and funding | 8 |
| 29 | Describe plans, strategies, or suggestions to evaluate impact | Integration in decision-making, funding allocation, review of relevant documents | N/A |
| J | Funding and conflict of interest |  |  |
| 30 | State sources of funding | Name sources of funding for the priority-setting exercise; if relevant include the budget and/or cost | 15 |
| 31 | Declare any conflicts or competing interests | State any conflicts of interest that may be at an individual level and/or at a contextual level (e.g. political issues, controversies) that may affect the process, output or implementation. | 15 |
